# Performance and Robustness of Machine Learning-based Radiomic COVID-19 Severity Prediction

**DOI:** 10.1101/2020.09.07.20189977

**Authors:** Stephen S.F. Yip, Zan Klanecek, Shotaro Naganawa, John Kim, Andrej Studen, Luciano Rivetti, Robert Jeraj

## Abstract

**Objectives:** This study investigated the performance and robustness of radiomics in predicting COVID-19 severity in a large public cohort.

**Methods:** A public dataset of 1110 COVID-19 patients (1 CT/patient) was used. Using CTs and clinical data, each patient was classified into mild, moderate, and severe by two observers: (1) dataset provider and (2) a board-certified radiologist. For each CT, 107 radiomic features were extracted. The dataset was randomly divided into a training (60%) and holdout validation (40%) set. During training, features were selected and combined into a logistic regression model for predicting severe cases from mild and moderate cases. The models were trained and validated on the classifications by both observers. AUC quantified the predictive power of models. To determine model robustness, the trained models was cross-validated on the inter-observer’s classifications.

**Results:** A single feature alone was sufficient to predict *mild from severe* COVID-19 with 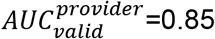 and 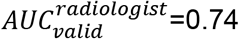 (p<< 0.01). The most predictive features were the distribution of small size-zones (GLSZM-SmallAreaEmphasis) for provider’s classification and linear dependency of neighboring voxels (GLCM-Correlation) for radiologist’s classification. Cross-validation showed that both 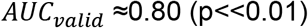. In predicting *moderate from severe COVID-19*, first-order-Median alone had sufficient predictive power of 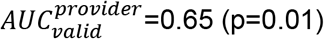. For radiologist’s classification, the predictive power of the model increased to 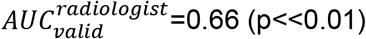 as the number of features grew from 1 to 5. Cross-validation yielded 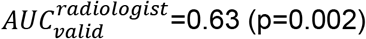 and 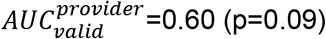.

**Conclusions:** Radiomics significantly predicted different levels of COVID-19 severity. The prediction was moderately sensitive to inter-observer classifications, and thus need to be used with caution.

**Key points:**

- Interpretable radiomic features can predict different levels of COVID-19 severity
- Machine Learning-based radiomic models were moderately sensitive to inter-observer classifications, and thus need to be used with caution

## INTRODUCTION

Coronavirus disease 2019 (COVID-19) pandemic has not only posed great threat to global health, but also put tremendous burden on the healthcare system [1, 2]. Accurate prediction of COVID-19 severity could provide actionable insights in guiding crucial hospitalization and treatment decisions, thereby alleviating the healthcare system strain [3].

Chest computed tomography (CT) is an invaluable tool for evaluating pulmonary involvement of COVID-19 [4–7]. Common qualitative CT imaging features of COVID-19 pneumonia include bilateral and peripheral ground glass opacities (GGO) distribution with/without consolidation [4]. As the disease progresses, total lung involvement increases along with the presence of crazy-paving and reverse halo sign [4, 6]. Thus, the radiologist’s manual assessment of these CT-based features has demonstrated great promise in COVID-19 diagnosis [5, 8, 9], prognosis prediction [10], and determining disease severity and therapeutic response [6, 11, 12]. However, manual assessment is laborious and subtle CT imaging findings can be overlooked by radiologists, especially due to the increased workload amid the outbreak. Accurate quantification of distinct COVID-19 imaging phenotypes may help automate the detection of sophisticated imaging features for disease severity prediction, reducing radiologists’ workload. Furthermore, quantification would allow for more accurate assessment of COVID-19 treatment response for treatment development support. One way to quantify such phenotypes is to employ radiomics [13].

Radiomics automatically computes an atlas of complex features for medical image phenotypic characterization by utilizing mathematical algorithms that quantify relationships among image voxels [13]. Radiomic features have been used to quantify disease or organ phenotypes for predicting clinical outcome [14, 15], treatment response [16, 17], genetic alteration [18, 19], and severity of pulmonary injury [20–23]. Recent studies by Wei et al (2020) [23] and Homayounieh et al [24] have demonstrated great promise in using radiomic features to predict COVID-19 severity based on classifications manually assessed by radiologists. However, the effect of inter-radiologist classification on the severity prediction was not investigated by the studies [5, 24, 25]. Further, they used both > 1000 unfiltered and filtered (e.g. wavelet) -based radiomic features [26, 27]. Filtered features are difficult to intuitively interpret [27]. As our understanding of COVID-19 is constantly evolving, use of filtered features may lack flexibility for incorporating new knowledge and troubleshooting when outliers occur.

In this study, we employed interpretable lung radiomic features to predict different levels of COVID-19 severity in a large public cohort of 1110 patients and investigated the robustness of the prediction with respect to inter-observer classifications. To our knowledge, this is the largest COVID-19 radiomic study.

## METHODS AND MATERIALS

### Patient population

This study used a publicly available MosMedData dataset (**https://mosmed.ai**) provided by municipal hospitals in Moscow, Russia from March—April, 2020 [25]. This dataset consists of chest CT images of 1110 COVID-19 patients (1 CT/patient) and were classified into different levels of lung tissue damage severity (Table 1). The population is 42% male and has a median age of 47 years (18—97 years). Our research workflow is shown in Fig 1.

**Figure 1.**
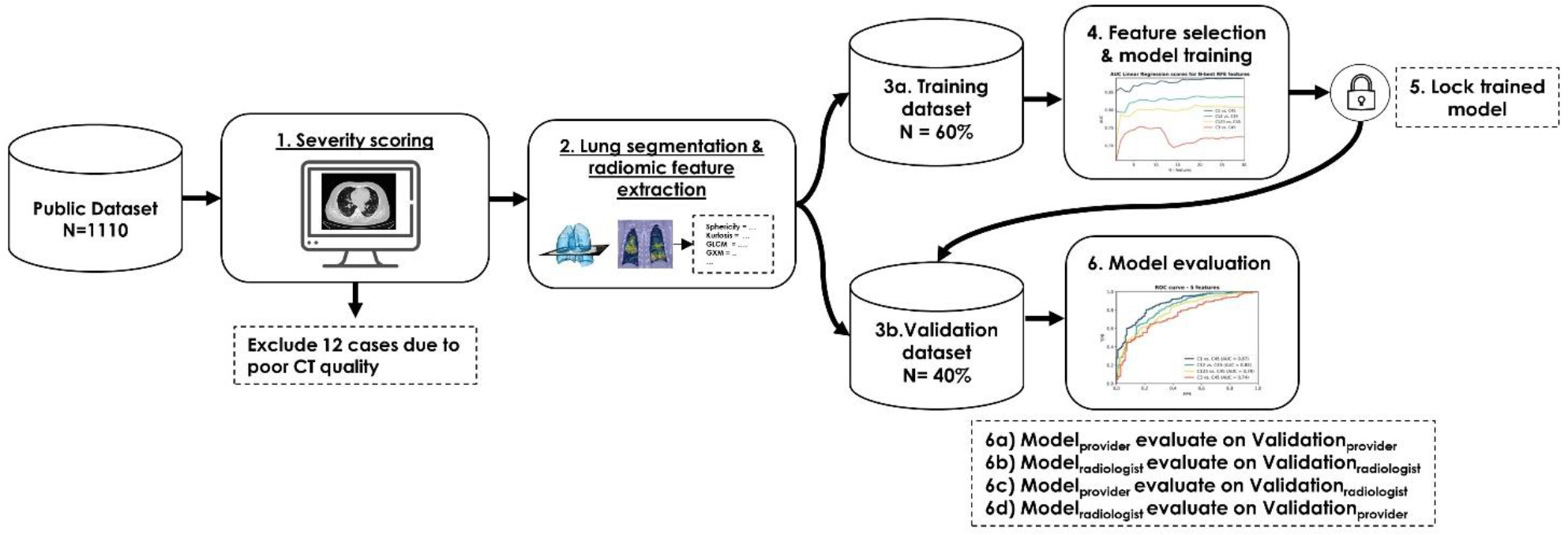
Research workflow: 1) Each patient was assigned with a CO-RADS score. 2) Lung mask was segmented by the watershed algorithm and radiomic features were extracted within the mask. 3) Dataset was then randomly divided into training and holdout validation dataset. 4) Feature selection and model training were performed. 5) Trained model was then locked and applied to validation data for 6) model evaluation.

**Table 1.**
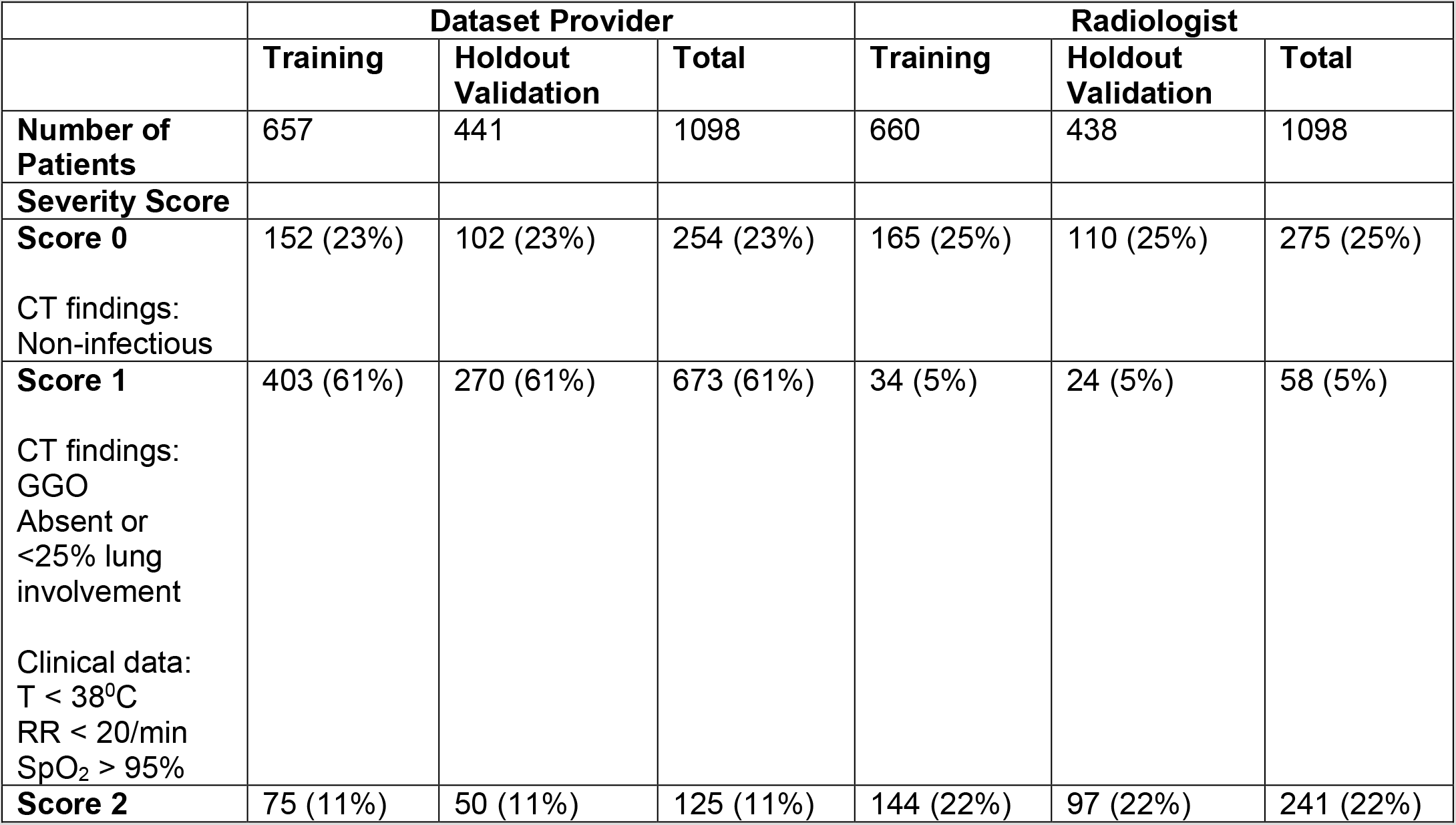
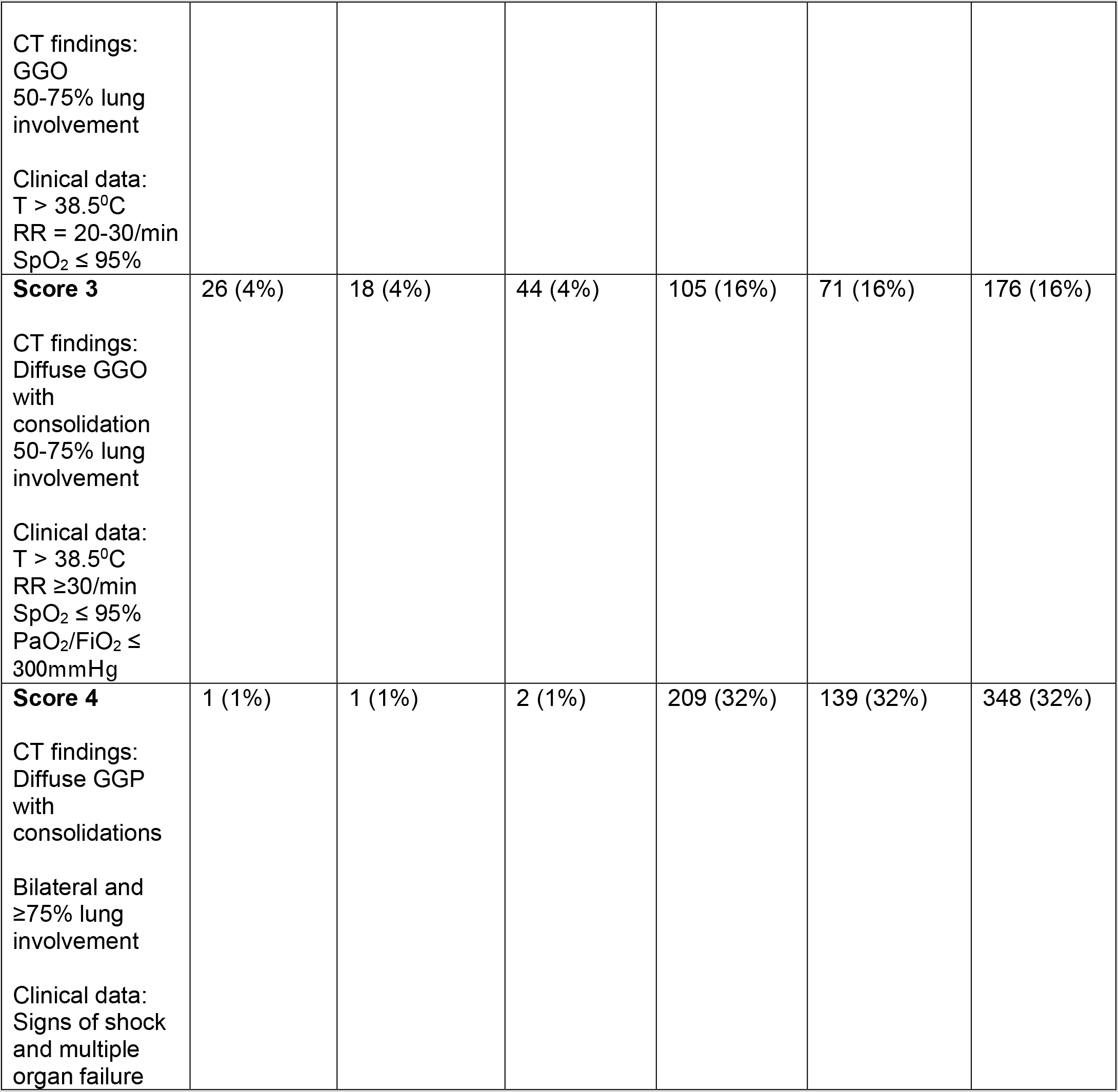
Severity score determined by data provider and a radiologist. T = temperature. RR = respiratory rate. SpO_2_ = peripheral capillary oxygen saturation. PaO_2_ = partial pressure of oxygen. FiO_2_ = Fraction of inspired oxygen. GGO = ground glass opacities.

|  | Dataset Provider |  |  | Radiologist |  |  |
| --- | --- | --- | --- | --- | --- | --- |
|  | Training | Holdout Validation | Total | Training | Holdout Validation | Total |
| **Number of Patients** | 657 | 441 | 1098 | 660 | 438 | 1098 |
| **Severity Score** |  |  |  |  |  |  |
| **Score 0**<br>CT findings:<br>Non-infectious | 152 (23%) | 102 (23%) | 254 (23%) | 165 (25%) | 110 (25%) | 275 (25%) |
| **Score 1**<br>CT findings:<br>GGO<br>Absent or<br><25% lung<br>involvement<br>Clinical data:<br>T < 38°C<br>RR < 20/min<br>SpO<sub>2</sub> > 95% | 403 (61%) | 270 (61%) | 673 (61%) | 34 (5%) | 24 (5%) | 58 (5%) |
| **Score 2** | 75 (11%) | 50 (11%) | 125 (11%) | 144 (22%) | 97 (22%) | 241 (22%) |
| CT findings:<br>GGO<br>50-75% lung involvement<br>Clinical data:<br>T > 38.5°C<br>RR = 20-30/min<br>SpO<sub>2</sub> ≤ 95% |  |  |  |  |  |  |
| **Score 3**<br>CT findings:<br>Diffuse GGO with consolidation<br>50-75% lung involvement<br>Clinical data:<br>T > 38.5°C<br>RR ≥30/min<br>SpO<sub>2</sub> ≤ 95%<br>PaO<sub>2</sub>/FiO<sub>2</sub> ≤ 300mmHg | 26 (4%) | 18 (4%) | 44 (4%) | 105 (16%) | 71 (16%) | 176 (16%) |
| **Score 4**<br>CT findings:<br>Diffuse GGP with consolidations<br>Bilateral and ≥75% lung involvement<br>Clinical data:<br>Signs of shock and multiple organ failure | 1 (1%) | 1 (1%) | 2 (1%) | 209 (32%) | 139 (32%) | 348 (32%) |

### COVID-19 Severity Classification

Using CT findings and clinical data (Table 1), each patient was assigned a severity score range 0 (very mild) to 4 (critical) by two inter-observers: (1) the dataset provider and (2) our board-certified radiologist with 7 years of chest CT interpretation experience The scoring criteria and distribution among patients are shown in Table 1, respectively. Further, CTs of twelve patients with substantial motion artifacts were excluded from further analysis.

### Lung segmentation and radiomic feature extraction

Lung masks were segmented using watershed segmentation algorithm [28] implemented in lung contouring function of AIQ Solutions’ Pulmonary Solutions technology. The watershed algorithm was initialized based on CT images with the threshold of -200HU. All segmented lung masks were then reviewed and manually adjusted by either an independent radiologist or an experienced imaging scientist.

A total of 107 non-filtered radiomic features were extracted within the segmented lung from the CT images using Python’s PyRadiomics v.3.0 package [26, 29]. The extracted features included 18 first-order, 14 shape, 24 gray level co-occurrence matrix (GLCM), 16 gray level run length matrix, 16 gray level size zone matrix (GLSZM), 5 neighboring gray tone difference matrix (NGTDM), and 14 gray level dependence matrix (GLDM) features. The extracted features are listed in the Supplementary Table 1. Equation for each feature is described in [26, 29]. Prior to radiomic feature extraction, the voxel size of all CT images and their lung segmentations were resampled to uniform size of 3×3×3mm^3^[13, 30]. The density of CT images was also discretized using a fixed width of 25HU to increase computation efficiency [13, 26]. Only non-filtered features were used due to their interpretability (Table 2). Analysis of 1116 LoG and wavelet filtered features are shown in the Supplementary Fig 1.

**Table 2.** List of top ranked features for both classification problems (i.e. Mild vs Severe COVID-19 and Moderate vs Severe COVID-19) that were considered for machine learning models. Provider = Dataset Provider’s classification. Rad = Radiologist’s classification. GLCM = Gray level co-occurrence matrix, GLDM = Gray level dependence matrix, GLSZM = Gray level size zone matrix, and NGTDM = Neighboring gray tone difference matrix

| Radiomic Type | Radiomic feature | Mild vs Severe | Moderate vs Severe | Description |
| --- | --- | --- | --- | --- |
| **First-order** |  |  |  |  |
|  | Median |  | Provider | Median value of the density distribution |
|  | 90percentile |  | Rad | 10th percentile of the density distribution |
|  | MeanAbsoluteDeviation |  | Rad | Mean distance of all density values from the average distance value |
|  | Mean |  | Rad | Average value of the density distribution |
| **GLCM** |  |  |  |  |
|  | Correlation | Rad | Rad | Measure the linear dependency of neighboring voxel density |
|  | InformationalMeasureofCorrelation2 |  | Rad | Measure correlation of the density distributions of neighboring voxels based on mutual information |
| **GLSZM** |  |  |  | A zone defined as 3D volume with similar density value |
|  | SmallAreaEmphasis | Provider |  | Quantify distribution of small size zones. High value indicates fine and heterogeneous textures |

### Statistical analysis

Two statistical analyses (classification problems) were conducted to investigate the ability of radiomic features in predicting different levels of COVID-19 severity:

1. mild vs severe (score 0 and 1 vs score 3 and 4)
2. moderate vs severe (score 2 vs score 3 and 4)

The patients with very mild (score 0) and mild (score 1) were grouped into mild severity category. Additionally, patients with severe (score 3) and critical (score 4) suspicion were grouped into severe category.

The predictive power was quantified by the area under the receiver operating characteristics curve (AUC). We used a permutation test to investigate if AUC was significantly different from random guessing (AUC = 0.50). In the permutation test, classifications were randomized 100,000 times and AUC was computed for each randomization. AUC was considered significant when fewer than 5000 AUCs computed from randomization (p< 0.05 = 5000/100,000) ≥ AUC of interest [31].

### Radiomic-based machine learning model training and validation

The dataset was randomly divided into training (60%) and holdout validation (40%) dataset (Table 1). During training, maximum relevance minimum redundancy (MRMR) algorithm and recursive feature elimination (RFE) method were used for radiomic feature selection implemented in MATLAB fscmrmr function and Python’s Scikit-learn, respectively [32]. Specifically, features were ranked according to their relationship with the prediction class (relevance) and among themselves (redundancy). Both relationships were quantified using mutual information [32]. With RFE, the best 30 MRMR filtered features were added to a machine learning (ML) logistic regression model.

The model was applied to the training set for training and to conduct the statistical analyses. The set of 30 best MRMR features was pruned recursively based on five-fold cross validation. Average training AUC (AUC_train_) and its standard deviation of the model were computed for each pruning. The set of features with the highest average AUC_train_ was selected. The ML model with the selected features was trained on the entire training set and locked (i.e. no further training) and then applied to the holdout validation set. Validation AUC (AUC_valid_) was computed to evaluate the predictive power of the trained radiomic-based ML model.

All aforementioned analyses were conducted using both provider- 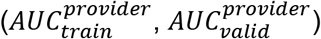 and radiologist- 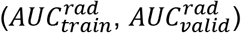 determined classification.

### Robustness Study

Radiomic-based ML models that were trained based on the training dataset of provider-determined classifications was validated on the hold out validation dataset of the radiologist-determined classifications 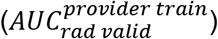, and vice versa. The models were considered robust if AUC_valid_ is significantly different from 0.50 (random guessing). Robustness study’s AUCs were indicated by 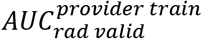 and 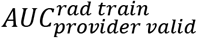.

### Classification Agreement

Cohen’s Kappa statistics (K) was used to quantify the agreement between provider- and radiologist-determined classification. Cohen’s K ranges from −1 to 1 with |K|≤0.10 = poor, 0.10<|K|≤0.20 = slight, 0.20<|K|≤0.40 = fair, 0.40<|K|≤0.60 = moderate, 0.60<|K|≤0.80 = substantial, |K|>0.80 = excellent agreement.

## RESULTS

This study investigated the ability of radiomic features in predicting different levels of COVID-19 severity in a large patient cohort. Visual examples of patients with different severity are shown in Fig 2.

**Figure 2.**
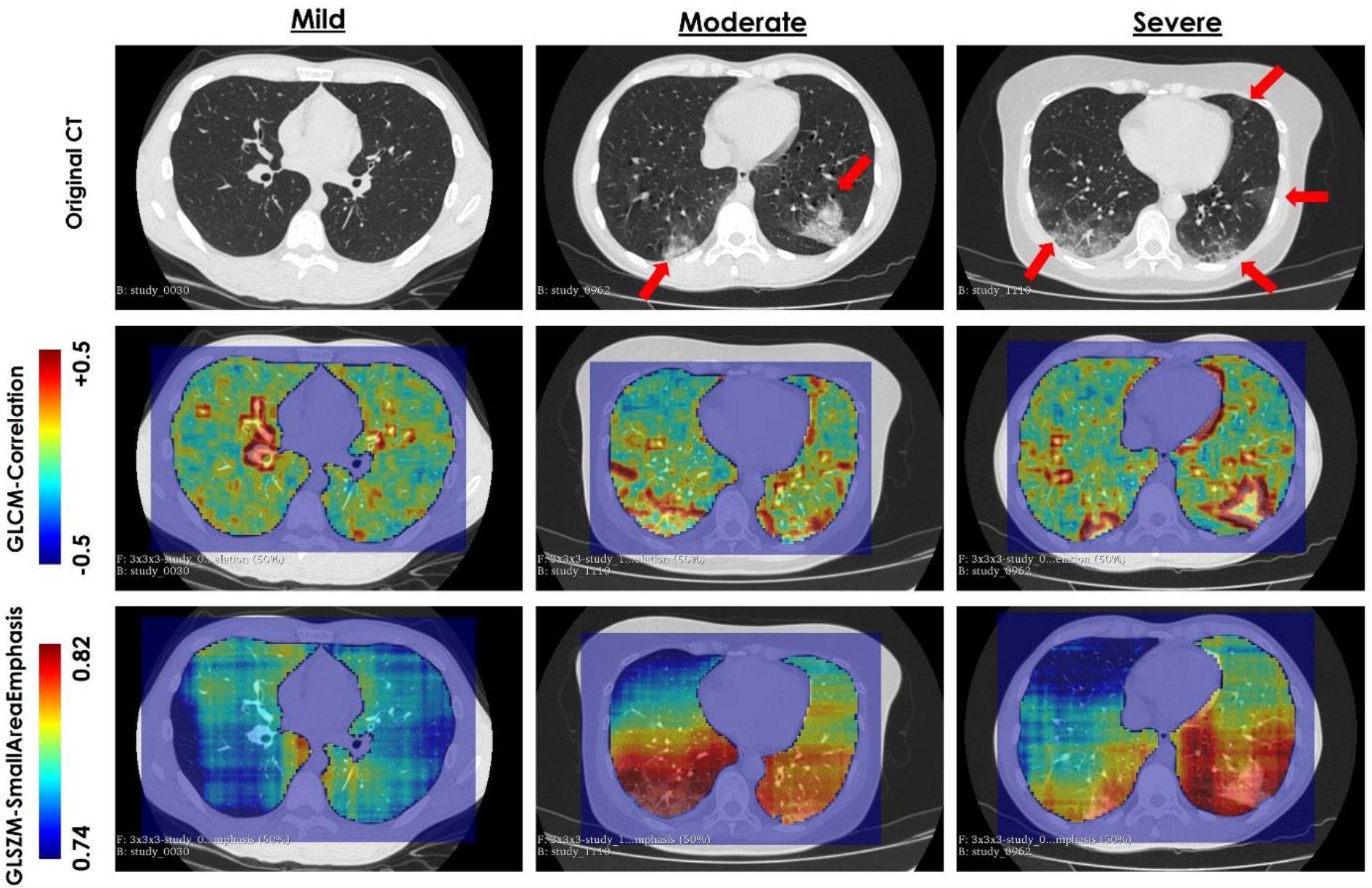
Radiomic filtered images. The mild, moderate, and severe COVID-19 patients were both identified by the data provider and theradiologist. Figure 3a) – c) are an axial slice of original CT display in lung window with W = 1400HU and L = –500HU. Red arrows indicate location of infections. Figure 3d) – f) are GLCM-correlation filtered images overlaid on CTs. Figure 3g) – i) are the GLSZM-SmallAreaEmphasis filtered images overlaid on CTs. GLCM = Gray level co-occurrence matrix, GLDM = Gray level dependence matrix, GLSZM = Gray level size zone matrix

**Figure 3.**
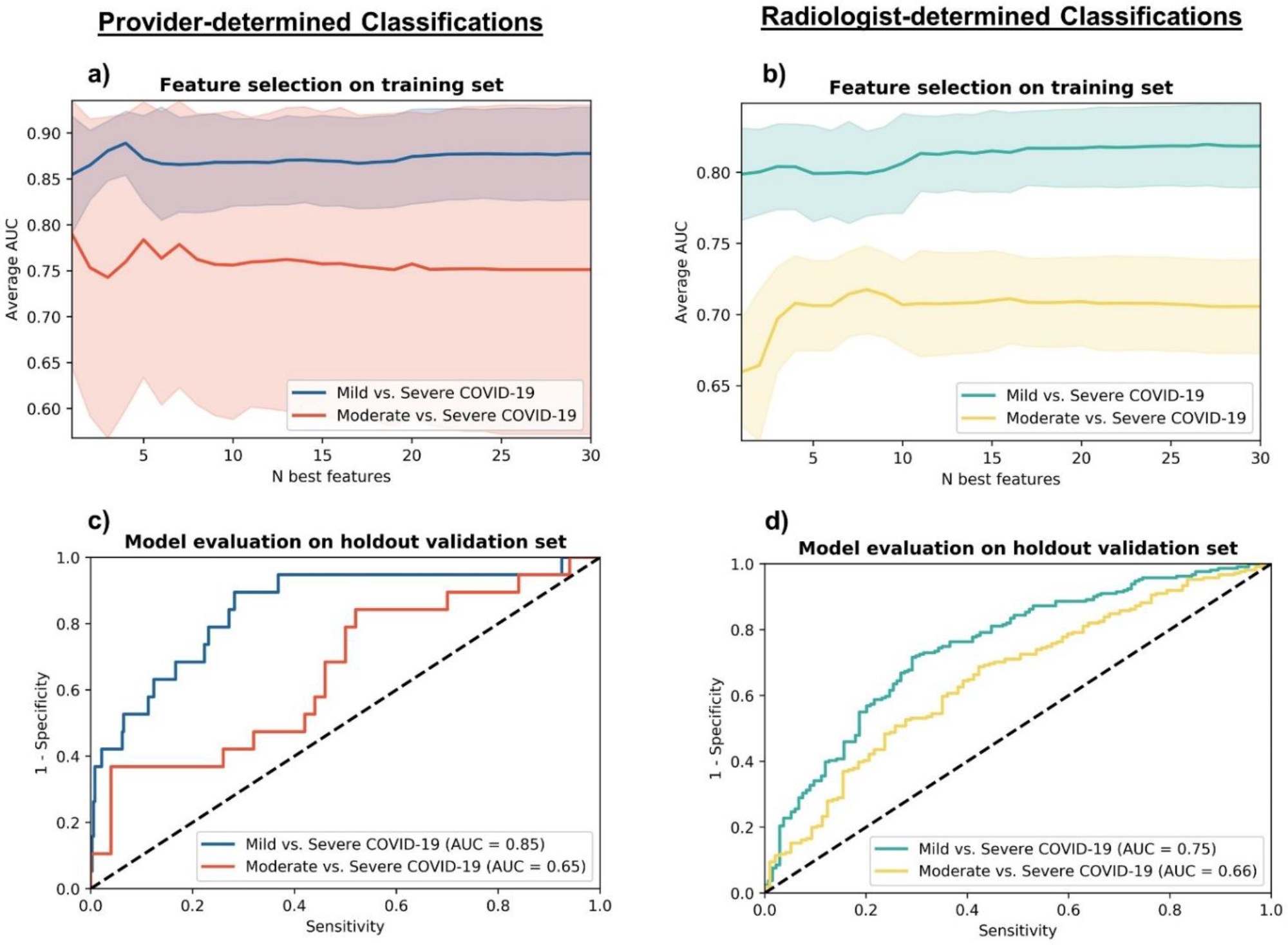
Results of radiomic model selection and training for (a) the provider’s and (b) the radiologist’ classifications, where shaded areas are the standard deviation of 5-cross validation. Model evaluation on holdout validation dataset for (c) the provider’s and (d) the radiologist’s classification.

### Mild vs Severe COVID-19

In training, as the number of features in our ML model was pruned from 5 to 1, average AUC_train_ changed by < 2% (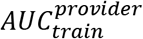 range: 0.85±0.06 and 0.87±0.05 and 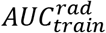 range: 0.80±0.03 and 0.80±0.03) (Fig 2a-b). As observed in Fig 2, more than one feature did not improve ML model performance. Therefore, only a single GLSZM-SmallAreaEmphasis and GLCM-Correlation feature was selected for provider’s and radiologist’s classification model validation, respectively (Fig 2c-d). In validation, univariate analysis also showed that GLSZM-SmallAreaEmphasis significantly predict mild from severe COVID-19 cases with 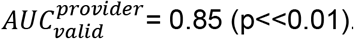. GLCM-Correlation separated mild from severe cases with 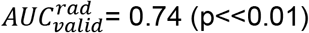 (Fig 2d).

The ML model trained on the provider-determined classifications resulted in 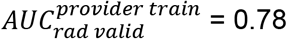 (p<< 0.01) (Fig 3a) in separating mild from severe COVID-19 cases, while the radiologist-determined classifications trained model had 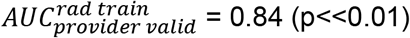 (Fig 3b).

### Moderate vs Severe COVID-19

In training, adding more than one feature in the ML model did not improve average 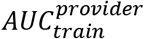 (Fig 2a). For example, combining top 5 MRMR ranked features (i.e. (1) first-order-Median, (2) GLCM-Correlation, (3) first-order-90percentile, (4) first-order-RootMeanSquared, and (5) first-order-Mean changed for average 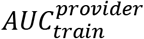 from 0.79±0.15 to 0.78±0.15. In validation, univariate analysis showed that first-order-Median significantly differentiated moderate from severe class with 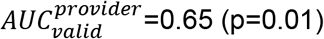 (Fig 2c).

On the other hand, the average 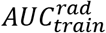 of the ML model increased from 0.65±0.04 to 0.70±0.03 as the number of features grew from 1 to 5 (Fig 2b). The top 5 MRMR ranked features for 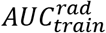 were: (1) GLCM-Correlation, (2) first-order-10percentile (3) GLCM-IMC2 (4) first-order-MeanAbsoluteDeviation and (5) first-order-Median (Table 2). More than 5 features did not further improve 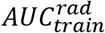 as observed in Fig 2b. All 5 features were selected for validation. In validation, ML logistic regression model based on these top 5 features yielded 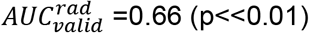 (Fig 2d).

The ML model trained on the provider-determined classifications had 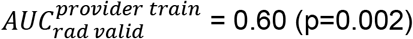 (Fig 3a) in moderate and severe cases prediction, while ML model trained the radiologist-determined classifications had 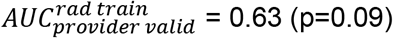 (Fig 3b).

### Classification Agreement

The Cohen’s |K| for mild, moderate, and severe class was 0.13, 0.06, and 0.06, respectively. |K| for all three classes was 0.10.

## DISCUSSION

Chest CT plays an important role in COVID-19 management [6]. COVID-19 induced pulmonary injury can exhibit a number of observable CT imaging phenotypes [4]. Our study investigated the performance and robustness of automatically computed radiomic features in quantifying distinct COVID-19 imaging phenotypes for disease severity prediction in a large patient cohort.

Our study has demonstrated that a single or few radiomic features can predict different levels of COVID-19 severity. In mild and severe COVID-19 separation, the predictive power of the ML models remained high (AUC_valid_>0.70) and significant (p<< 0.01) regardless of being trained on the provider- or radiologist-determined classifications (Fig 2). On the other hand, ML models had a modest performance of AUCvalidation∼0.65 in moderate from severe cases prediction (Fig 2). Although the radiologist’s classification trained ML model had 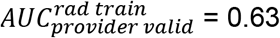 on the provider’s hold out validation dataset, the prediction was not significant, suggesting moderate sensitivity of radiomic-based ML models to the inter-observer classification (Fig 3b). Radiomic-based ML models thus should be used with caution and as an ML-assisted severity assessment tools. Although radiomics has great potential to improve radiologists’ workflow amid the COVID-19 outbreak, the model outputs should be reviewed and signed off by an experienced radiologist.

Our Cohen’s Kappa analysis suggested that there was only a small agreement between the provider-and radiologist-determined classifications. Manual scoring systems, such as BI-RADS and Lung-RADS scores for cancer screening, often suffered from substantial inter-observer discordance [33]. To reduce disagreement, some studies [19, 27] included more than two radiologists and used the majority vote to determine the final score. If all radiologists scored differently, they would review both clinical and imaging data together and discuss any discrepancies until the consensus was achieved [19, 27]. To improve the robustness of radiomic-based ML models in the COVID-19 severity prediction, the models should be trained on classifications based on the unanimous decision arrived from three or more inter-observers.

Interestingly, the radiomic model trained by Wei et al (2020) [23] achieved an excellent performance with an AUC of 0.93 while the performance of our models were lower (Fig 3 and 4). Additionally, including additional 1116 wavelet and LoG radiomic features did not improve our model performance (Supplementary Fig 1). The difference in model performance between our and the study by Wei et al [23] may be due to the patient size (1110 patients in our study vs only 81 patients in the study by Wei et al. [23]) and race (Russian vs Chinese cohort). It would be interesting to also investigate if COVID-19 could drive distinct pulmonary injury patterns among different races. Further, Homayounieh et al (2020) [24] also employed CT radiomics to predict COVID-19 severity in 315 patients from Iran. However, their study used hospital admission and survival data as surrogate for COVID-19 severity while our study predicted severity classifications subjectively determined by experts. Future studies with external dataset of CT images, hospitalization, and outcome data should be used to validate the radiomics-based imaging biomarkers identified in our study.

**Figure 4.**
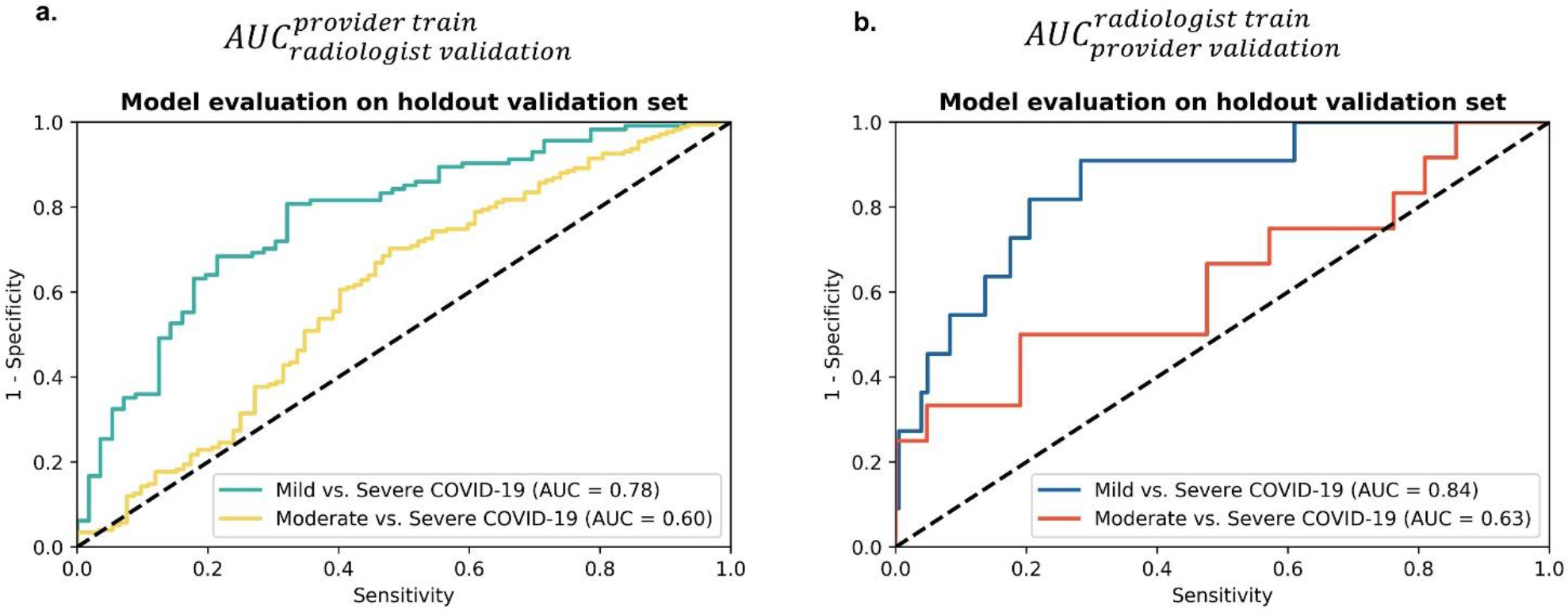
Cross Validation. (a) Models trained on the provider’s classifications was validated on the hold out dataset of the radiologist’s classifications. (b) Models trained on the radiologist’s classifications was validated on the hold out dataset of the provider’s classifications.

Lungs with high degree of HU histogram spread and nonuniform texture are likely to be COVID-19 infected. In severity classification, we observed substantial GGO distribution and consolidation giving rise to image contrast between normal lung and infected tissues, and heterogeneous lung appearance (Fig 2). While normal lung tissue has HU unit of <-700, COVID-19 induced GGO and consolidation have much higher CT density [37, 38]. Thus, increased pulmonary involvement in severe cases can lead to high CT density values. This explains why radiomic features (e.g. first-order Median and 10Percentile) that quantify overall CT density values played an important role in detecting patients with more severe COVID-19 (Table 2). Furthermore, in the radiomic filtered images shown in Fig 2, GLCM-correlation was observed to be high around pulmonary vessels (Fig 2d-e) and in the infected areas (Figure 2f), but low in the normal pulmonary tissues (Fig 2d-f). This observation suggests that although the voxels of normal lung are similar, they tend to be less linearly dependent. In addition, large pulmonary involvement of COVID-19 patients could also lead to greater overall GLCM-correlation values. This may explain why GLCM-correlation alone could significantly separate mild from severe COVID-19 patients. Moreover, similar GLCM-correlation patterns were observed in both the moderate and severe COVID-19 patient, implying that, as expected, moderate and COVID-19 infected patients could share similar imaging characteristics (Fig 2e-f). Further, since GLSZM-SmallAreaEmphasis captures both the heterogeneity of histogram distribution and variations in zone sizes (i.e. extensions of infection) (Table 2), it also helped significantly differentiate mild from severe cases.

Our use of descriptive radiomic features provide transparency into what drove the prediction of ML model (e.g. heterogeneous texture and spread of density distribution). Using only handful of features, radiomics demonstrated significant ability to separate patients into different severity levels of COVID-19 infection on a validation set (Fig 3 and 4). Although deep learning models may identify latent imaging patterns for accurate COVID-19 detection [39, 40], they are difficult to intuitively interpret and thus often regarded as a “black-box” [27, 41]. Deep learning algorithms usually have > 1 million parameters [42]. As our understanding of COVID-19 is constantly evolving, such black box approach lacks flexibility for incorporating new knowledge and troubleshooting when outliers occur. However, with careful integration of an ensemble model of radiologist-defined, radiomic, and deep learning features one could further improve COVID-19 diagnosis while also efficiently incorporating new knowledge.

## CONCLUSION

Radiomic-based ML models predicted patients with different levels of COVID-19 severity, particularly in the mild and severe case separation. However, inter-observer classifications modestly affected moderate and severe cases prediction. Our study suggests that radiomics may be useful for early identifying severe COVID-19 cases for hospital admission or treatment management but need to be used with cautions.

## Data Availability

This public dataset is freely available for scientific research on https://mosmed.ai

## ACKNOWLEDGMENTS

The authors would like to thank the Center of Diagnostics and Telemedicine in Moscow, Russia for making the MosMedData COVID19 Dataset publicly available.

## Abbreviations

COVID-19: Coronavirus disease 2019
GLCM: Gray level co-occurrence matrix
GLDM: Gray level dependence matrix
GLSZM: Gray level size zone matrix
IMC2: Informational measure of correlation 2
ML: Machine learning
MRMR: Maximum relevance and minimum redundancy
RFE: Recursive feature elimination

## Notes

### Competing Interest Statement

S.S.F.Y. and R.J. of this manuscript declare relationships with AIQ Solutions Inc: S.S.F.Y. is an employee and a shareholder of AIQ Solutions, Inc. R.J. is a consultant and shareholder of AIQ Solutions, Inc.

### Funding Statement

The authors state that this work has not received any funding.

### Author Declarations

Institutional Review Board approval was not required because public MosMedData dataset was used. This public dataset is freely available for scientific research on https://mosmed.ai

